# Hospitalisations and deaths due to Ambulatory Care Sensitive Conditions (ACSC) among adults with and without Intellectual Disabilities in Scotland

**DOI:** 10.1101/2024.02.22.24303205

**Authors:** Filip Sosenko, Deborah Cairns, Bhautesh D Jani, Laura Ward, Maria Truesdale, Laura Hughes-McCormack, Angela Henderson, Craig Melville

## Abstract

**Background:** Conditions that should be sufficiently managed in primary health care are collectively known as Ambulatory Care Sensitive Conditions (ACSC). The rate of unplanned hospital admissions for ACSC can be regarded as a proxy indicator of how well the primary care system works for a population of interest. We investigate such rates in Scotland, focusing on adults with Intellectual Disabilities (ID) and contrasting them with adults without ID.

**Method:** A population-based retrospective cohort data linkage study of adult respondents to Scotland’s 2011 Census. Self- or proxy-reported ID status from the Census was linked to hospital admissions data and deaths data. The cohort was followed until the end of 2019.

**Results:** After adjusting for different ACSC prevalence in ID and non-ID cohorts, we did not find evidence of there being a higher risk of unplanned ACSC hospitalisation among people with ID. COPD, seizures and epilepsy, influenza and pneumonia were responsible for half of ACSC hospitalisations, regardless of ID status. However, adults with ID had a higher risk of dying due to ACSC than adults without ID.

**Conclusions:** We conclude that overall, the primary care system in Scotland appears to be similarly effective for adults with ID than for adults without ID. However, the higher risk of dying from ACSC among people with ID needs further research.

## Introduction

Adults with Intellectual Disabilities (ID) often experience complex and multiple health needs [1–3] which are different to those experienced by adults in the general population [2]. Frequency of hospital admissions for adults with ID has also been reported as higher [4, 5] and evidence suggests that adults with ID face numerous barriers to managing long-term conditions in primary care due to: communication difficulties; partial health literacy; difficulty understanding health information; and patients feeling fear or embarrassment [6, 7]. Barriers are also created through lack of education/knowledge about people with ID from healthcare professionals; inadequate coordination and continuity of care; and limited involvement of people with ID and supporters/carers in healthcare decision-making [6, 8–10]. Due to the existence of these barriers, annual health checks and other such active interventions are being introduced for adults with ID [11].

Since some disadvantaged groups in the population, including people with ID, have more limited access to / engagement with primary care, or may receive less effective care, some researchers have argued that the rate of unplanned hospital admissions for Ambulatory Care Sensitive Conditions (ACSC)(such as asthma or diabetes) can be used as a proxy indicator of how well the primary care system works for a population of interest [12–17] [18]. Thus, a higher the rate of unplanned hospital admissions ACSC is taken as an indicator of a less effective primary care system.,

A number of empirical studies, particularly in the US, have shown that in countries where access to primary health care is not universal, people experiencing socio-economic disadvantage (who thus have more limited access) have a higher rate of unplanned hospitalisations for ACSC [19, 20][21, 22]. However, some studies have not found any such link [21, 23, 24]. A counter-argument has also been raised that it is not obvious that unplanned hospitalisations that occur are truly preventable [25]. It has also been pointed out that primary care providers should not be held responsible for unplanned hospitalisations where the subjects did not seek medical care [21].

With regards to substantive findings about ACSC among people with ID, previous research in the UK found that such people were consistently hospitalised at a higher rate for ACSC than people without an ID [26, 27]. However, neither of these studies controlled for the prevalence of ACSC in the analyses. A US study on diabetes and asthma, did control for different prevalences of and reported that the hospitalisation rates for diabetes and asthma were higher for adults with ID [28]. Another US study found that ACSC hospitalisation rates were higher for women than men with ID [29]. A study in England found that health checks in primary care reduced emergency admissions for ACSC [30], while a US study has found that people residing at home with no health support services had a higher emergency hospital visit rate than people with other residential circumstances [31].

In this paper we take a view that unplanned hospitalisation rates for ACSC are a useful indicator of the extent to which there is a ‘problem’ when it comes to access to primary care, or the quality of care received, for a specific group cf. the general population. We do agree, however, with an argument made by some researchers (e.g. [21]) that the rate needs to be adjusted for the prevalence of a given condition in each population being compared. It is logical to expect more ACSC hospitalisations in the population with a higher prevalence of ACSC. However, all proxy measures, hospitalisation rates for ACSC lacks nuance as an indicator of the quality of primary care services. There is also an additional element when considering the effectiveness of primary care for adults with ID because the initial presentation to primary care services is often dependent on family or paid carers recognising there is a problem. Nonetheless, our view is that ACSC hospitalisation rate gives an overall indication of the effectiveness of the reactive primary care system for adults with ID [32].

The main aim of this study was to investigate whether the ACSC hospitalisation rate among people with ID in Scotland is higher than the general population rate. An additional aim was to compare mortality due to ACSC, a topic that has received relatively less attention so far [18, 33, 34]

## Methods

### Study design

The research was a population-based cohort data linkage study of people who took part in Scotland’s 2011 Census and were at least 18 years old on census day (27 March 2011). The study was focused on adults only because health conditions often have different prevalence in children and adults, and the primary care system in Scotland (as in many other countries) is arranged somewhat differently for children than adults. Therefore including children in the study would have required a separate analysis, which was beyond the scope of the current project.

The two study cohorts consisted of all adults with ID and a 15% randomly selected comparator sample from the general population. The follow-up period ended on 31 December 2019. Census data was linked to individual-level data about hospitalisations and deaths.

The ID population included those with co-occurring autism while the general population cohort excluded people with ID or autism. The individual’s ID status was determined from census data; one of the questions regarded disabilities and the list included ‘learning disability’ (the term much more widely used in the UK than ‘intellectual disability’, but with the same meaning). Census information was provided by a proxy where the respondent was not capable of completing the form. People who self- or proxy-identified as from the general population but who subsequently died of ID (ICD-10 codes F70, F71, F72, F73, F78, F79, F84) were excluded. People who died on the census day were also excluded. Unplanned ACSC hospitalisations were identified in hospital data using a codelist and specification provided by NHS Digital [35] (Supplementary File 1).

The study was approved by Scotland’s Statistics Public Benefit and Privacy Panel (reference: 1819-0051) and the University of Glasgow College of Medical, Veterinary and Life Sciences Ethical Committee.

### Data

The three datasets used for this project (Scotland’s 2011 Census, hospital admissions, death certificate data) were provisioned by Electronic Data Research and Innovation Service (eDRIS, part of Public Health Scotland) via the Scottish National Safe Haven, a platform for researchers working with large databases of electronic health records^1^. The hospital admission dataset (SMR01) is described in detail in the National Data Catalogue^2^. The death certificate data is maintained by the National Records for Scotland. The datasets were merged by the authors using unique anonymised personal identifiers provided by eDRIS.

The prevalence of specific ACSC among the ID population in Scotland is unknown, and we did not have access to primary care data from which it could be estimated. We therefore used data from England, specifically the National Health Service (NHS) data series ‘Health and Care of People with Learning Disabilities’^3^. We regard the Scottish and English ID populations to be close enough in character for it to not be an issue for our analysis. While only six of 19 ACSC conditions are covered by this data source, collectively they are responsible for over half of all ACSC hospitalisations in Scotland (see below).

### Data analyses

#### Unplanned ACSC hospitalisations

The exposure was ID status, and the outcome was unplanned hospital admission that met the NHS-provided specification requirements for one of the ACSC [35]. Note that while most ACSC diagnoses in this specification are based on a given ACSC being recorded as ‘primary’ diagnosis, five of the 19 ACSC (Influenza and pneumonia, other vaccine preventable, Chronic Obstructive Pulmonary Disease (COPD), diabetes complications, gangrene) can meet the specification when they are recorded as non-primary diagnosis. For this reason, there is no ground for making a distinction between ‘hospitalisation due to ACSC’ and ‘hospitalisation with ACSC’ based on the diagnostic position.

The analysis focused on incidence rate ratios, both crude and age-sex adjusted. We also report cumulative incidences as these are easier to understand for audiences without background in epidemiology. Additionally, they may be more useful for evaluating the burden on the health care system, as for this it does not matter whether two hospitalisations come from the same person or from two persons. (This matters in the calculation of time at risk for rates, though). However, as people with ID live on average shorter than people without ID [36], and obviously people who leave the cohort cannot be hospitalised, incidence rates are generally a more appropriate measure in this context than cumulative incidences.

#### Deaths due to ACSC and deaths involving ACSC

The analysis of mortality was somewhat different to the analysis of unplanned hospitalisations. While primarily interested in rate ratios, it was informative to investigate what proportion of all deaths in each population were due to ACSC and to distinguish between ‘deaths due to ACSC’, where ACSC is reported as main-cause of death on the death certificate and from ‘deaths involving ACSC’, where ACSC is reported in any position on the death certificate (‘all-cause’).

In all analyses statistical significance was defined at the conventional 5% level and determined by examining the overlap of 95% Confidence intervals. The analyses were conducted in Stata 17. The code is available from the authors upon request. Additional tables for specific age groups (18-54, 55+, 18-64, 65+) and for men and women are also available on request.

## Results

### Characteristics of the study population

The size of the whole adult ID population on census day was 16,840. The size of the 15% general population random sample was 566,074 adults. The total number of person-years was 4,818,262, of which 134,769 in the ID cohort and 4,683,493 in the general population.

Adults with ID were on average younger than adults without ID (Table 1). The ID population had relatively more males than females in comparison to the general population. On average, people with ID tended to live in somewhat more deprived areas than people without ID.

**Table 1.**
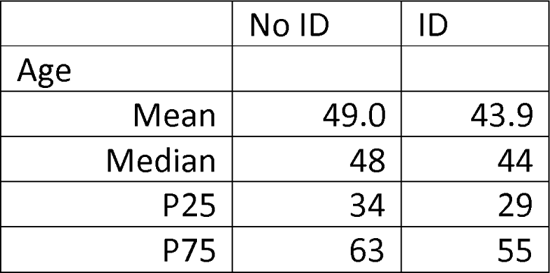
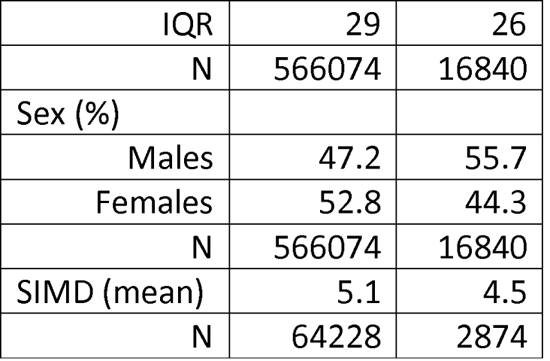
Age, sex and area deprivation profile of adults on Census Day, by ID status.

|  | No ID | ID |
| --- | --- | --- |
| Age |  |  |
| Mean | 49.0 | 43.9 |
| Median | 48 | 44 |
| P25 | 34 | 29 |
| P75 | 63 | 55 |
| IQR | 29 | 26 |
| N | 566074 | 16840 |
| Sex (%) |  |  |
| Males | 47.2 | 55.7 |
| Females | 52.8 | 44.3 |
| N | 566074 | 16840 |
| SIMD (mean) | 5.1 | 4.5 |
| N | 64228 | 2874 |

### Unplanned ACSC hospitalisations

#### Cumulative incidence

The cumulative incidence of unplanned hospitalisations for ACSC was overall similar for the ID and general population. Around 5% of all hospitalisations were due to ACSC, regardless of ID status (Table A1). Around 4% of unique persons had one or more ACSC hospitalisation, again regardless of ACSC status (Table 2). Seizures and epilepsy, COPD and influenza and pneumonia were responsible for half of all ACSC hospitalisations among people without ID, and for 55% of all ACSC among people with ID.

**Table 2.**
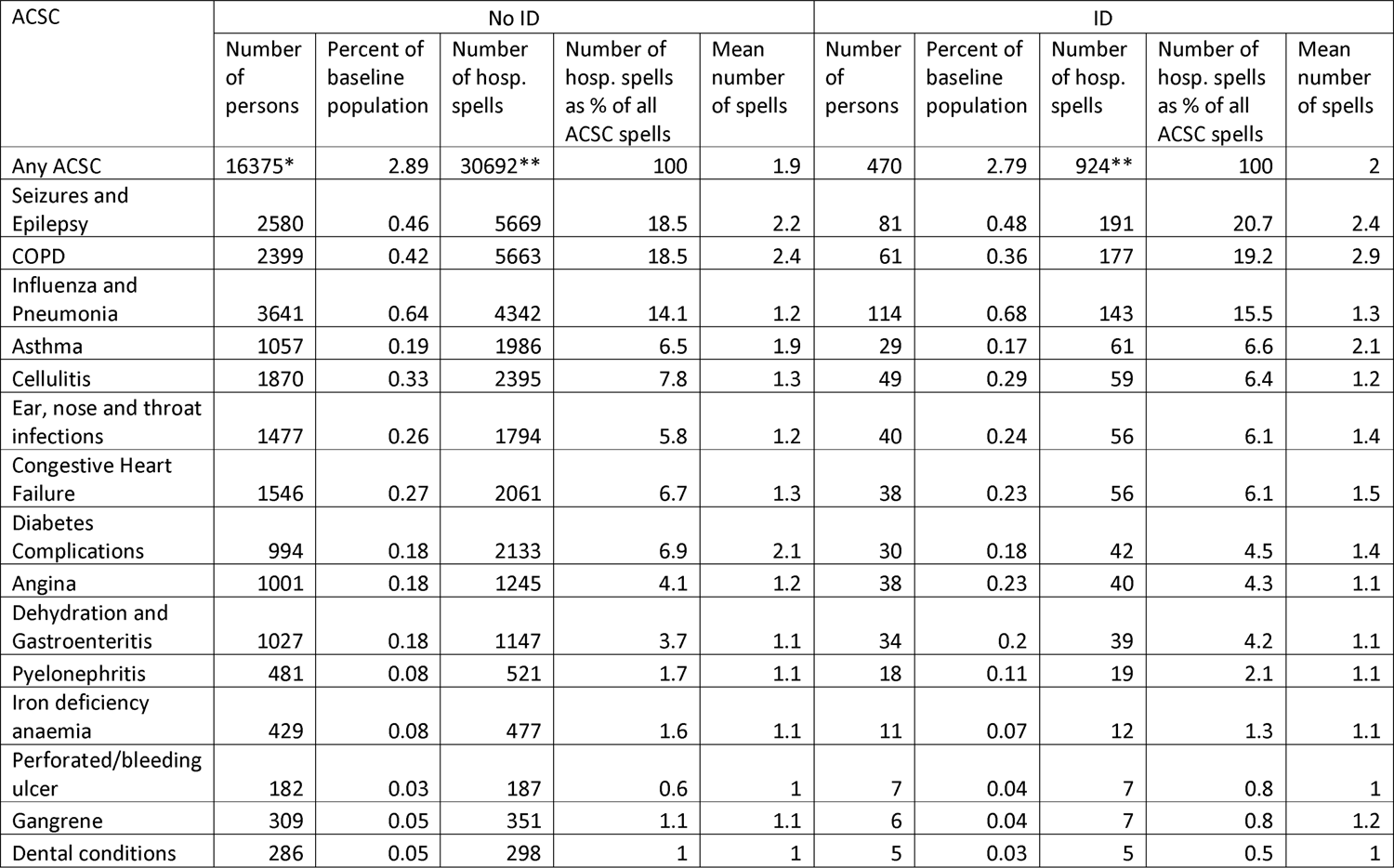
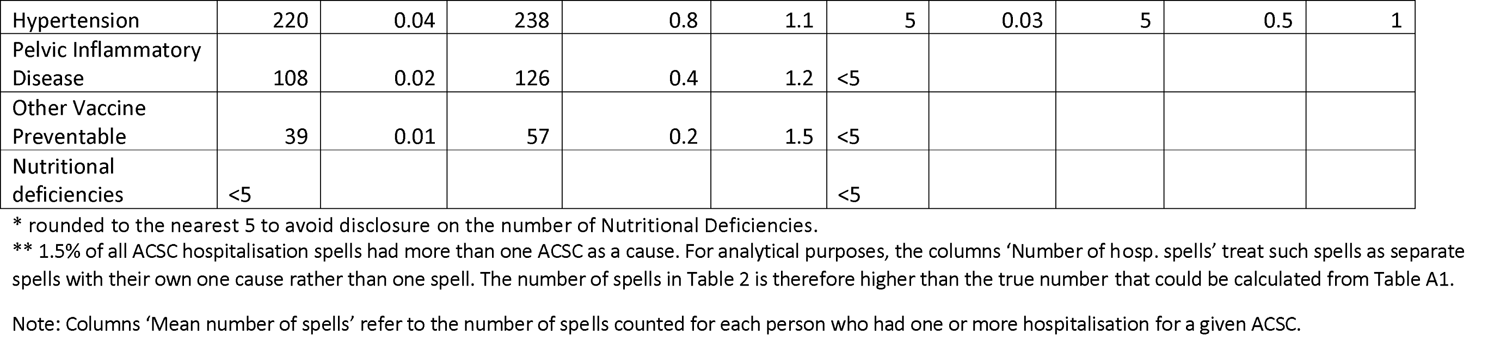
Statistics on adults alive on census day who had at least one unplanned hospitalisation due to ACSC during the study period, by ID status and ACSC, 2011-19 (sorted by decreasing order of ‘Number of hosp. spells’ in ID)

| ACSC | No ID |  |  |  |  | ID |  |  |  |  |
| --- | --- | --- | --- | --- | --- | --- | --- | --- | --- | --- |
|  | Number of persons | Percent of baseline population | Number of hosp. spells | Number of hosp. spells as % of all ACSC spells | Mean number of spells | Number of persons | Percent of baseline population | Number of hosp. spells | Number of hosp. spells as % of all ACSC spells | Mean number of spells |
| Any ACSC | 16375* | 2.89 | 30692** | 100 | 1.9 | 470 | 2.79 | 924** | 100 | 2 |
| Seizures and Epilepsy | 2580 | 0.46 | 5669 | 18.5 | 2.2 | 81 | 0.48 | 191 | 20.7 | 2.4 |
| COPD | 2399 | 0.42 | 5663 | 18.5 | 2.4 | 61 | 0.36 | 177 | 19.2 | 2.9 |
| Influenza and Pneumonia | 3641 | 0.64 | 4342 | 14.1 | 1.2 | 114 | 0.68 | 143 | 15.5 | 1.3 |
| Asthma | 1057 | 0.19 | 1986 | 6.5 | 1.9 | 29 | 0.17 | 61 | 6.6 | 2.1 |
| Cellulitis | 1870 | 0.33 | 2395 | 7.8 | 1.3 | 49 | 0.29 | 59 | 6.4 | 1.2 |
| Ear, nose and throat infections | 1477 | 0.26 | 1794 | 5.8 | 1.2 | 40 | 0.24 | 56 | 6.1 | 1.4 |
| Congestive Heart Failure | 1546 | 0.27 | 2061 | 6.7 | 1.3 | 38 | 0.23 | 56 | 6.1 | 1.5 |
| Diabetes Complications | 994 | 0.18 | 2133 | 6.9 | 2.1 | 30 | 0.18 | 42 | 4.5 | 1.4 |
| Angina | 1001 | 0.18 | 1245 | 4.1 | 1.2 | 38 | 0.23 | 40 | 4.3 | 1.1 |
| Dehydration and Gastroenteritis | 1027 | 0.18 | 1147 | 3.7 | 1.1 | 34 | 0.2 | 39 | 4.2 | 1.1 |
| Pyelonephritis | 481 | 0.08 | 521 | 1.7 | 1.1 | 18 | 0.11 | 19 | 2.1 | 1.1 |
| Iron deficiency anaemia | 429 | 0.08 | 477 | 1.6 | 1.1 | 11 | 0.07 | 12 | 1.3 | 1.1 |
| Perforated/bleeding ulcer | 182 | 0.03 | 187 | 0.6 | 1 | 7 | 0.04 | 7 | 0.8 | 1 |
| Gangrene | 309 | 0.05 | 351 | 1.1 | 1.1 | 6 | 0.04 | 7 | 0.8 | 1.2 |
| Dental conditions | 286 | 0.05 | 298 | 1 | 1 | 5 | 0.03 | 5 | 0.5 | 1 |
| Hypertension | 220 | 0.04 | 238 | 0.8 | 1.1 | 5 | 0.03 | 5 | 0.5 | 1 |
| Pelvic Inflammatory Disease | 108 | 0.02 | 126 | 0.4 | 1.2 | <5 |  |  |  |  |
| Other Vaccine Preventable | 39 | 0.01 | 57 | 0.2 | 1.5 | <5 |  |  |  |  |
| Nutritional deficiencies | <5 |  |  |  |  | <5 |  |  |  |  |

\* rounded to the nearest 5 to avoid disclosure on the number of Nutritional Deficiencies.
\*\* 1.5% of all ACSC hospitalisation spells had more than one ACSC as a cause. For analytical purposes, the columns 'Number of hosp. spells' treat such spells as separate spells with their own one cause rather than one spell. The number of spells in Table 2 is therefore higher than the true number that could be calculated from Table A1.
Note: Columns 'Mean number of spells' refer to the number of spells counted for each person who had one or more hospitalisation for a given ACSC.

Those who had at least one ACSC hospitalisation had on average just over 2 ACSC spells in the follow-up period, in both populations (Table 2). COPD was the ACSC with the highest mean number of spells per person. Overall means were similar in both cohorts, except for diabetes complications which had a noticeably lower mean in the ID cohort (1.4 vs 2.1). The average length of hospital stay was similar between the two cohorts (Table A3).

#### Incidence rates and ratios

The crude incidence rates of unplanned hospitalisations for ‘any ACSC’ were similar in both ID and general populations, with the difference not being statistically significant (Table A2). The standardized incidence rate for ID was similar to the crude rate.

As would be expected from cumulative incidence findings, seizures and epilepsy, COPD and influenza and pneumonia had the highest incidence rates in both cohorts, and incidence rates were similar between people with ID and the general population.

Most incidence rate ratios were close to 1.0 (Table 3), which is unsurprising in the light of the fact that most incidence rates were similar in both populations. Only one SIR was statistically significantly different from 1.0: in the case of diabetes complications, it was lower than 1.0.

**Table 3.** Ratios of incidence rates of adult hospitalisations due to an ACSC per 1,000 person-years, ID population (nominator) compared with No ID population (denominator), 2011-19 (sorted by decreasing SIR)

| ACSC | IRR | IRR 95% CI | SIR | SIR 95% CI |
| --- | --- | --- | --- | --- |
| Any ACSC | 1.01 | (0.94, 1.07) | 0.98 | (0.91, 1.05) |
| Perforated/bleeding ulcer | 1.26u | (0.50, 2.64)u | 1.44 | (0.66, 3.14) |
| Pyelonephritis | 1.22u | (0.73, 1.93)u | 1.18 | (0.74, 1.90) |
| seizures and Epilepsy | 1.14 | (0.98, 1.31) | 1.17 | (1.00, 1.36) |
| Influenza and Pneumonia | 1.11 | (0.93, 1.31) | 1.09 | (0.91, 1.30) |
| Dehydration and Gastroenteritis | 1.14 | (0.81, 1.57) | 1.07 | (0.77, 1.49) |
| COPD | 1.05 | (0.90, 1.22) | 1.03 | (0.88, 1.21) |
| Ear, nose and throat infections | 1.06 | (0.79, 1.38) | 1.01 | (0.76, 1.35) |
| Asthma | 1.03 | (0.79, 1.33) | 0.95 | (0.73, 1.23) |
| Angina | 1.08 | (0.77, 1.48) | 0.94 | (0.68, 1.29) |
| Cellulitis | 0.83 | (0.63, 1.07) | 0.86 | (0.65, 1.15) |
| Congestive Heart Failure | 0.91 | (0.69, 1.19) | 0.84 | (0.64, 1.11) |
| Hypertension | 0.71u | (0.23, 1.68)u | 0.75 | (0.31, 1.83) |
| Iron deficiency anaemia | 0.84u | (0.43, 1.49)u | 0.72 | (0.40, 1.30) |
| Gangrene | 0.67u | (0.27, 1.40)u | 0.61 | (0.28, 1.31) |
| Diabetes Complications | 0.66 | (0.48, 0.90) | 0.59 | (0.43, 0.81) |
| Dental conditions | 0.57u | (0.18, 1.34)u | 0.53 | (0.21, 1.29) |
| Other Vaccine Preventable |  |  |  |  |
| Nutritional deficiencies |  |  |  |  |
| Pelvic Inflammatory Disease |  |  |  |  |

Note: letter 'u' indicates that the figure may not be reliable due to the small number of cases (between 5-20).

When the populations were broken down by age bands and sex, some more patterns started to appear. Among adults under 65, the SIR for seizures and epilepsy was statistically significantly above 1.0 while SIR for asthma was significantly above 1.0 among adults under 55 (see the Appendix). SIR for congestive heart failure was significantly below 1.0 among people aged 55+. SIR for influenza and pneumonia was significantly above 1.0 among adult women under 55 and SIR for COPD was significantly above 1.0 among adult men under 65.

As reported earlier, it is more appropriate to consider incidence rate ratios not by themselves but in the context of population prevalence. Table 4 presents prevalence ratios for six ACSC. Source: Health and Care of People with Learning Disabilities data series^4^. Point estimates and 95% Confidence Intervals (CI) were calculated by the authors.

**Table 4.** Crude and standardized prevalence ratios, ID adults (nominator) compared with no-ID adults (denominator), by ACSC, 2021-22, England.

| ACSC | PR | PR 95% CI | Age-sex standardized PR (SPR) | 95% CI for age-sex standardized PR (SPR) |
| --- | --- | --- | --- | --- |
| COPD | 0.66 | (0.63, 0.69) | 0.96 | (0.91, 1.01) |
| Asthma | 1.54 | (1.52, 1.57) | 1.58 | (1.55, 1.61) |
| Convulsions and Epilepsy | 24.01 | (23.71, 24.31) | 28.09 | (27.74, 28.45) |
| Congestive Heart Failure | 0.98 | (0.93, 1.02) | 1.4 | (1.33, 1.48) |
| Hypertension | 0.74 | (0.73, 0.75) | 1.01 | (1.00, 1.03) |
| Diabetes* | 1.42 | (1.39, 1.44) | 1.78 | (1.75, 1.81) |
| *Non-type 1 Diabetes* | 1.39 | (1.37, 1.42) | 1.79 | (1.76, 1.82) |
| *Type 1 Diabetes* | 1.79 | (1.69, 1.90) | 1.67 | (1.57, 1.77) |

\* To allow for comparisons with diabetes complications in our hospitalisations and deaths data, figures for diabetes have been calculated by us by aggregating non-type 1 and type 1 diabetes. This is not ideal as non-type 1 and type 1 can co-occur. However, this happens relatively rarely [37].
Source: Health and Care of People with Learning Disabilities data series<sup>4</sup>. Point estimates and 95% Confidence Intervals (CI) were calculated by the authors.

Taking prevalence into consideration, the SIR for hospitalisations was statistically significantly lower than the SPR (the upper bound of 95% CI for SIR was lower than the lower bound of 95% CI for SPR) in the case of asthma (part. men), convulsions and epilepsy, congestive heart failure and diabetes complications among all adults. Additionally, in the 18-54 age range, SIR was lower than SPR for congestive heart failure and COPD.

### Deaths among adults

#### Cumulative mortality

The proportion of all adults alive on Census day who died during the study was 11.4% in the No ID cohort and 16.7% in the ID cohort. With regards to ACSC ‘main-cause’, the proportions were 0.85% of the No ID and 1.3 % of the ID (Table 5). There were approximately 3.5 times more deaths involving ACSC (‘all-cause’): 3.2% of No ID adults and 4.5 % of adults in the ID cohort.

**Table 5.**
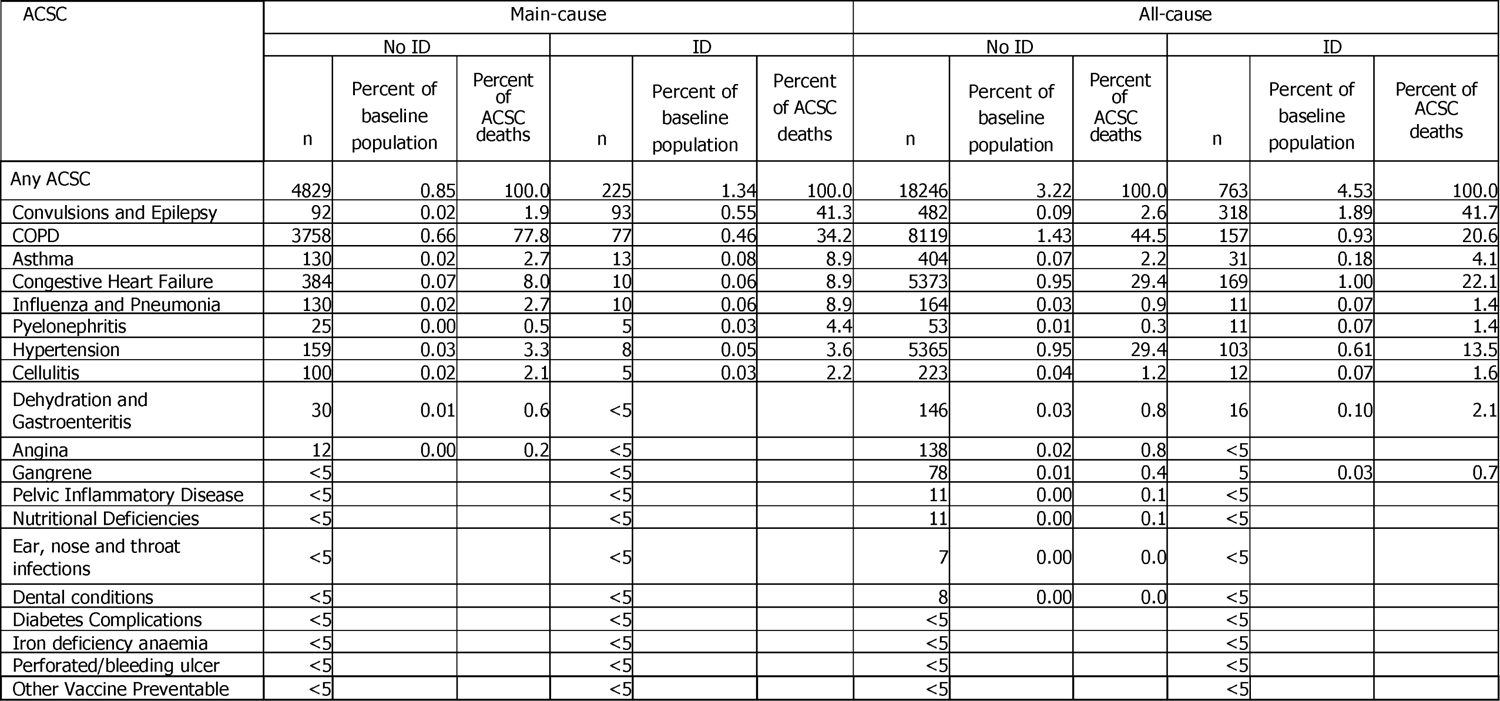
Number and percent of adults alive on census day who died during the study period and whose death certificate mentions ACSC as ‘main-cause of death’ or ‘all-cause of death’, by ID status, 2011-19 (sorted by decreasing order of ID main-cause n)

Of those adults who died during the study, 7.5% of people without ID died due to ACSC cf. 7.8% of people with ID. The proportion of all deaths that involved ACSC (all-cause) was 28.4% in the No ID cohort and 26.5% among people with ID. The mean age of adults at death was 77 in No ID cohort and 63 in the ID cohort. The mean age of adults at death due to ACSC was 78 in No ID and 62 in the ID cohort.

The vast majority (75%) of main-cause ACSC deaths in ID were caused by just two ACSCs: convulsions and epilepsy (41%) and COPD (34%) (Table 5). The picture was different among adults without ID, with COPD responsible for 78% of deaths and very few convulsions and epilepsy deaths (2%). Among women with ID, there were somewhat more ACSC deaths due to asthma, influenza and pneumonia and congestive heart failure than among men with ID (20% vs 8% of all ACSC deaths). Among older (55+) people with ID, COPD overtakes convulsions and epilepsy as the leading cause of ACSC deaths (45% vs 25%). Among older women (55+), the proportion of main-cause deaths due to asthma gradually increases to the point of becoming second-leading in the 65+ age group. With regards to all-cause ACSC deaths, apart from convulsions and epilepsy and COPD, congestive heart failure and hypertension are major all-cause causes of ACSC death, in both ID and non-ID populations.

### Mortality rates and ratios

People with ID had considerably higher ACSC mortality rates (both crude and age/sex-adjusted) than people without ID (Table A4). Without taking ACSC prevalence into account, main-cause Standardized Mortality Ratios (SMR) were statistically significantly higher on asthma (particularly among women), cellulitis, convulsions and epilepsy, COPD, hypertension, influenza and pneumonia (particularly among women), and pyelonephritis (particularly among women) (Table 6). Additionally, in the 18-64 age group the SMR on congestive heart failure was higher and statistically significant (particularly among women).

**Table 6.**
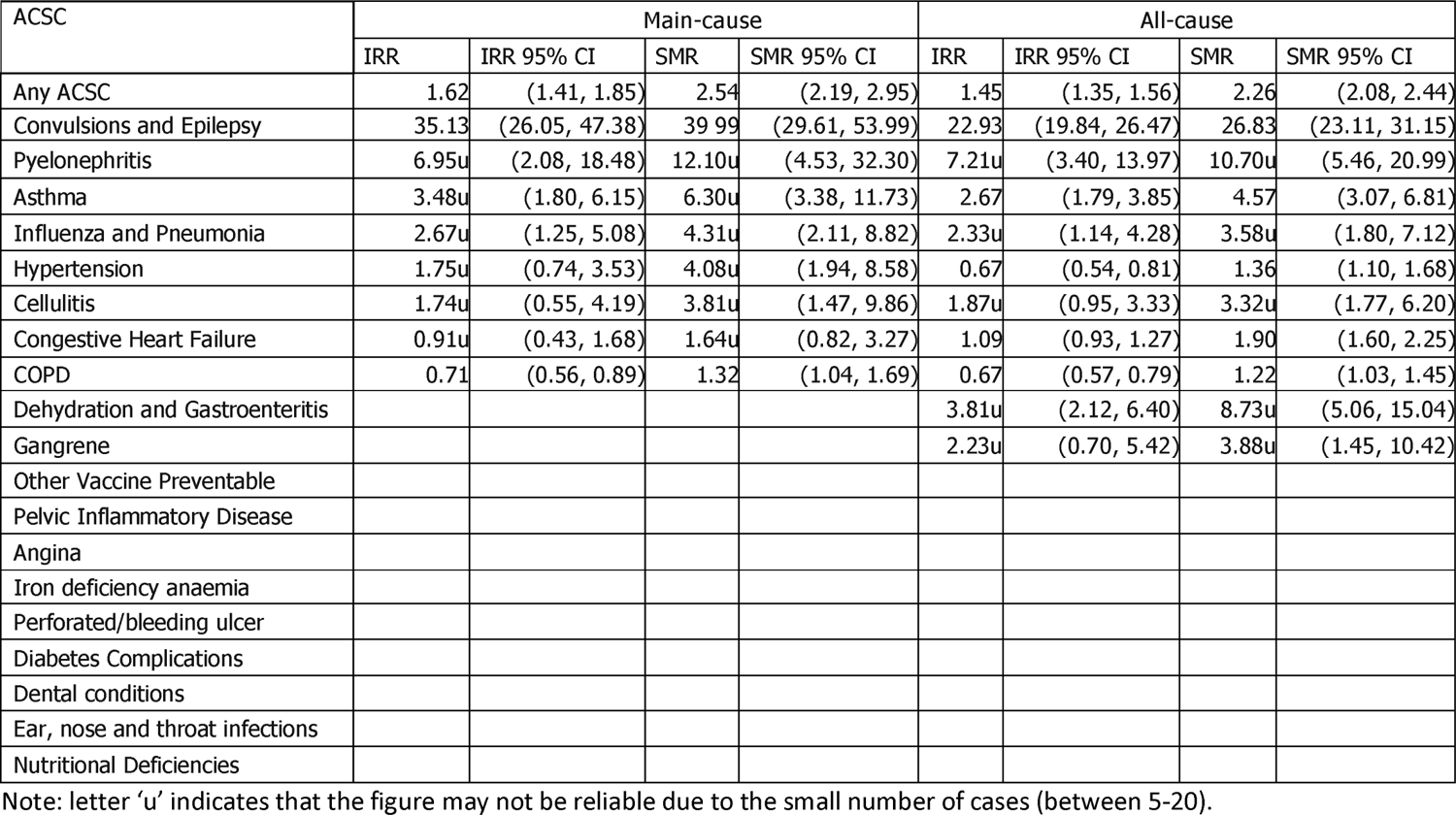
Adult ACSC mortality ratios by ‘main-cause of death’ status, ID cohort (nominator) compared with No ID cohort (denominator), 2011-19 (sorted by decreasing order of main-cause SMR)

When ACSC prevalence was taken into account, the above findings for asthma, convulsions and epilepsy, COPD, hypertension, and congestive heart failure remained unchanged. No prevalence data was available for cellulitis, influenza and pneumonia, and pyelonephritis.

Moving on to all-cause mortality, nominally SMRs were higher and statistically significant on asthma, cellulitis, convulsions and epilepsy, congestive heart failure, COPD, dehydration and gastroenteritis, gangrene, hypertension, influenza and pneumonia, and pyelonephritis. Taking prevalence into account, the findings were unchanged with regards to asthma, congestive heart failure, COPD and hypertension. SMR for convulsions and epilepsy was no longer statistically significant. No prevalence data was available for cellulitis, dehydration and gastroenteritis, gangrene, influenza and pneumonia, and pyelonephritis.

## Discussion

This is the first study to compare unplanned hospitalisations for ACSC and mortality from ACSC in adults with ID and adults who do not have ID. Although unplanned hospitalisations for ACSC were similar, the higher mortality rates for ACSC experienced by adults with ID is potentially important contributor to the inequalities in mortality, previously reported [36].

### Unplanned hospitalisations

It is clear that unplanned ACSC hospitalisations in Scotland constitute a non-negligible proportion of all hospitalisations (5%), regardless of ID status, burdening the health care budget in times of ever-increasing demand from an ageing population. It is also worth emphasising that people who have an unplanned ACSC hospitalisation are likely to have unplanned readmission to hospital for the same condition. This is consistent with existing evidence [38] and suggests room for targeted improvement in health care.

With regards to the question of which ACSC are leading causes of ACSC hospitalisations, both cumulative incidences and incidence rates show that COPD, seizures and epilepsy, and influenza and pneumonia are responsible for half of ACSC hospitalisations, in both populations.

As for what our findings tell us about the quality of primary care or the access to it, on their own, higher SIRs for convulsions and epilepsy, asthma, influenza and pneumonia, and COPD would suggest a negative story about how primary care in Scotland works for people with ID, with respect to those specific conditions. In contrast, the lower SIR on diabetes complications would suggest a positive story with regards to that specific condition. However, as argued in the introduction, it is more appropriate to consider incidence rate ratios not by themselves but in the context of population prevalence. When this is considered, not only the negative findings about seizures and epilepsy, asthma, influenza and pneumonia, and COPD disappear but the picture turns out to be positive: the hospitalisation ratio (SIR) is lower than the prevalence ratio (SPR); the difference is statistically significant as CIs do not overlap. Furthermore, the picture turns out to be positive in the 18-54 age range with regards to congestive heart failure. Therefore, in the case of these conditions as well as diabetes complications, the primary care system in Scotland seems to work at least as well for people with ID as for people without ID.

### Mortality

While the proportion of ACSC deaths in all deaths was similar in the two populations, our investigation of rates and ratios showed possible health inequalities for the population with ID. SMRs were higher than prevalence ratios for asthma, seizures and epilepsy, COPD, hypertension, and congestive heart failure. In line with existing evidence on shorter life expectancy among people with ID [36], the cumulative incidence of deaths in our study was much higher among people with ID. We did not have prevalence data about other ACSCs, so it is possible that other conditions have SMRs higher than prevalence ratios.

Since the vast majority of ACSC deaths in ID were caused by just two ACSCs: seizures and epilepsy and COPD, this implies that biggest gains in reducing mortality caused by ACSC could be achieved by targeting these two conditions. The ONS regards epilepsy as ‘treatable’ and COPD as ‘preventable’ [39].

Also, with regards to seizures and epilepsy, it is worth highlighting that it is one of the four leading causes of ACSC hospitalisations in the general population, but very few deaths are due to it (Table 5). Among people with ID, seizures and epilepsy produces both a lot of hospitalisations and a lot of deaths. This might suggest that when GPs treat an ID patient with seizures and epilepsy, they should be aware of this much higher risk of death.

### Understanding the findings

The study’s findings seem to be at tension: the picture is positive with regards to unplanned hospitalisations for ACSC and negative with regards to the increased mortality from ACSC experienced by adults with ID. The former suggests that the primary care system in Scotland works at least as well for people with ID as it does for the general population. However, the latter suggests that ACSCs may contribute to the inequalities in mortality experienced by adults with ID.

One clue to resolving this apparent contradiction is that in both populations almost no-one who died due to ACSC had a previous unplanned hospitalisation for that ACSC. If the rate of unplanned admission for ACSC is taken as an indicator of the quality of primary care, this suggests that adults with ID and adults who do not have ID are receiving a similar quality of primary care service. However, it could be that the severity of the ACSC is different at the time of the first hospitalisation for the ACSC. This is supported by the findings of a study that reported that at the time of hospitalisation, adults with ID have more severe pneumonia [40]. Greater disease severity at the time of hospitalisation could be due to the difficulties that adults with ID have seeking help for deteriorating health (Khoeiniha et al, 2021) and the problems health and social care staff have in recognising the deteriorating health status of adults with ID.

### Implications for policy, practice, and research

The study’s findings about ACSC hospitalisations seem to provide – at least at face value - a welcome exception from typically negative stories about the situation of people with ID. There are, however, four important qualifications that should be made. Firstly, the findings do not mean that the primary care system in Scotland is objectively ‘good’ for people with ID, they suggest it is overall ‘not worse’ for people with ID than for people without ID. The authors regularly meet people with ID and their carers (in research and use-of-services contexts), and their testimonies indicate that there is still significant room for improvement.

A second, related qualification regards a point made in the introduction, that using ACSC evidence from different countries presents an opportunity for policy or practice ‘transfer’. The ratio of ACSC hospitalisation rates to ACSC prevalence could be used to identify which countries can learn from which. Countries with a worse (higher) ratio than Scotland’s could benefit from transferring some Scottish policy and practice solutions. On the other hand, Scotland could benefit from looking into primary care systems in countries with a better (lower) ratio.

Thirdly, our findings about inequalities in ACSC mortality between people with ID and people without ID are not positive. More research is needed to understand why the picture of ACSC mortality is worse than the picture of ACSC hospitalisations.

Finally, it needs to be remembered that unplanned hospitalisations have a cost implication for the healthcare system, so even if people with ID are not (in relative terms) more likely to experience an ACSC hospitalisation than people without ID, it would still be beneficial to try to minimise the number of ACSC hospitalisations among people with ID.

### Strengths and limitations

The study’s main strength lies in the use of rigorous methodologies. The Census had a high response rate (94%) and the linkage of the Census to other datasets was carried out by a dedicated service (eDRIS) specialising in this kind of work, thus increasing our trust in the quality of the linkage. The dataset of hospital admissions and the dataset of death certificates are both long-standing and undergo high quality control. The list of ACSCs and the methodology for identification of ACSCs have been produced by the national healthcare provider, again increasing our confidence in the validity and reliability of the findings.

The study’s biggest limitation was the small size of the ID population in Scotland. This meant that our confidence intervals were often too wide to draw any conclusions. When confidence intervals are wide it is not possible to know whether differences in ratios reflect the reality or are just due to random chance.

Another limitation was that we had prevalence data only for six out of 19 ACSCs. However, within those six, two (convulsions and epilepsy, COPD) are the leading causes of ACSC hospitalisations in ID and four are the leading causes of all-cause mortality. Therefore, this limitation is relatively minor.

## Conclusions

The study has enhanced our knowledge and understanding of ACSC among people with ID in Scotland. It has established that after adjusting for disease prevalence, people with ID are at a lower risk of having an unplanned ACSC hospitalisation. Our conclusion is that the primary care system in Scotland works at least as well for people with ID as it does for the general population. This does not mean that there is room for complacency: 5% of all hospitalisations and 8% of deaths are due to ACSC and as such are potentially preventable. Large benefits in liveability and savings to the health care budget can be achieved by improvements in primary care.

## Data Availability

Data used in this study were available in the Scottish National Safe Haven (Project Number: 1819-0051), but as restrictions apply, they are not publicly available. Access to data may be granted on application to, and subject to approval by, the Public Benefit and Privacy Panel for Health and Social Care. Applications are coordinated by eDRIS (electronic Data Research and Innovation Service). The anonymised data used in this study was made available to accredited researchers only through the Public Health Scotland (PHS) eDRIS User agreement.

## Footnotes

1 https://www.publichealthscotland.scot/services/data-research-and-innovation-services/electronic-dataresearch-and-innovation-service-edris/national-safe-haven-nsh/

2 https://www.ndc.scot.nhs.uk/Data-Dictionary/SMR-Datasets/SMR01-General-Acute-Inpatient-and-Day-Case/

3 https://digital.nhs.uk/data-and-information/publications/statistical/health-and-care-of-people-with-learningdisabilities

4 https://digital.nhs.uk/data-and-information/publications/statistical/health-and-care-of-people-with-learning-disabilities

