## Appendix for "Hospitalisations and deaths due to Ambulatory Care Sensitive Conditions (ACSC) among adults with and without Intellectual Disabilities in Scotland"

Table A1. All adult hospitalisation spells* broken down by whether they were unplanned hospitalisations due to ACSC, by ID status, 2011-19 (%)

| | ACSC hospitalisation | | Total | N |
|---|---|---|---|---|
| | No | Yes | | |
| No ID | 94.8 | 5.2 | 100 | 579,917 |
| ID | 94.8 | 5.2 | 100 | 17,276 |

* A continuous inpatient (CIP) spell is a continuous period of care within the NHS, regardless of any transfers which may take place.

Table A2. Incidence rates of adult hospitalisations due to an ACSC per 1,000 person-years, by ID status and ACSC, 2011-19 (sorted by decreasing SIR in ID group)

| ACSC | Crude incidence rates | | | | ID incidence rates age-sex standardized to 'No ID' population | |
|---|---|---|---|---|---|---|
| | No ID | | ID | | | |
| | IR | IR 95% CI | IR | IR 95% CI | SIR | SIR 95% CI |
| Any ACSC | 6.07 | (6.00, 6.14) | 6.10 | (5.71, 6.51) | 5.94 | (5.56, 6.35) |
| Convulsions and Epilepsy | 1.17 | (1.14, 1.20) | 1.33 | (1.15, 1.53) | 1.36 | (1.18, 1.57) |
| COPD | 1.17 | (1.14, 1.20) | 1.23 | (1.06, 1.43) | 1.21 | (1.04, 1.40) |
| Influenza and Pneumonia | 0.90 | (0.87, 0.92) | 0.99 | (0.84, 1.17) | 0.98 | (0.83, 1.15) |
| Cellulitis | 0.50 | (0.48, 0.52) | 0.41 | (0.32, 0.53) | 0.43 | (0.33, 0.55) |
| Asthma | 0.41 | (0.39, 0.43) | 0.42 | (0.33, 0.55) | 0.39 | (0.30, 0.51) |
| Ear, nose, and throat inflections | 0.37 | (0.35, 0.39) | 0.39 | (0.30, 0.51) | 0.37 | (0.29, 0.49) |

| | | | | | | |
|---|---|---|---|---|---|---|
| Congestive Heart Failure | 0.43 | (0.41, 0.45) | 0.39 | (0.30, 0.51) | 0.36 | (0.27, 0.47) |
| Diabetes complications | 0.44 | (0.42, 0.46) | 0.29 | (0.22, 0.40) | 0.26 | (0.19, 0.36) |
| Dehydration and Gastroenteritis | 0.24 | (0.22, 0.25) | 0.27 | (0.20, 0.37) | 0.25 | (0.18, 0.35) |
| Angina | 0.26 | (0.24, 0.27) | 0.28 | (0.20, 0.38) | 0.24 | (0.17, 0.34) |
| Pyelonephritis | 0.11 | (0.10, 0.12) | 0.13u | (0.08, 0.21)u | 0.13u | (0.08, 0.20)u |
| Iron deficiency anaemia | 0.10 | (0.09, 0.11) | 0.08u | (0.05, 0.15)u | 0.07u | (0.04, 0.13)u |
| Perforated/bleeding ulcer | 0.04 | (0.03, 0.04) | 0.05u | (0.02, 0.10)u | 0.06u | (0.03, 0.11)u |
| Gangrene | 0.07 | (0.07, 0.08) | 0.05u | (0.02, 0.10)u | 0.04u | (0.02, 0.10)u |
| Hypertension | 0.05 | (0.04, 0.06) | 0.03u | (0.01, 0.08)u | 0.04u | (0.02, 0.09)u |
| Dental conditions | 0.06 | (0.05, 0.07) | 0.03u | (0.01, 0.08)u | 0.03u | (0.01, 0.08)u |
| Pelvic Inflammatory Disease | 0.03 | (0.02, 0.03) | | | | |
| Other Vaccine Preventable | 0.01 | (0.01, 0.02) | | | | |
| Nutritional deficiencies | | | | | | |

Note: letter 'u' indicates that the figure may not be reliable due to the small number of cases (between 5-20).

Table A3. Average length of unplanned hospital stay due to ACSC, by ID status and ACSC, 2011-19 (measured in days; stays of adults on Census 2011 day; length refers to hospital spells. Sorted by decreasing mean length of stay in the ID group)

| ACSC | No ID | | | ID | | |
|---|---|---|---|---|---|---|
| | Number of spells | median | mean | Number of spells | median | mean |
| Any ACSC | 30690* | 1.0 | 2.5 | 924 | 1.0 | 2.6 |

| | | | | | | |
|---|---|---|---|---|---|---|
| Dental conditions | 298 | 1.0 | 1.7 | 5 | 1.0 | 13.0 |
| Gangrene | 351 | 3.0 | 7.8 | 7 | 4.0 | 4.7 |
| Influenza and Pneumonia | 4342 | 1.0 | 3.2 | 143 | 1.0 | 3.8 |
| Cellulitis | 2395 | 1.0 | 3.1 | 59 | 1.0 | 3.7 |
| Perforated/bleeding ulcer | 187 | 1.0 | 2.4 | 7 | 2.0 | 3.4 |
| Diabetes complications | 2133 | 1.0 | 3.4 | 42 | 1.0 | 3.3 |
| COPD | 5663 | 1.0 | 2.5 | 177 | 1.0 | 2.9 |
| Congestive Heart Failure | 2061 | 1.0 | 3.3 | 56 | 1.0 | 2.4 |
| Dehydration and Gastroenteritis | 1147 | 1.0 | 3.2 | 39 | 1.0 | 2.1 |
| Pyelonephritis | 521 | 2.0 | 2.3 | 19 | 1.0 | 1.9 |
| Convulsions and Epilepsy | 5669 | 1.0 | 1.7 | 191 | 1.0 | 1.8 |
| Angina | 1245 | 1.0 | 1.3 | 40 | 1.0 | 1.6 |
| Ear, nose, and throat inflections | 1794 | 1.0 | 1.2 | 56 | 1.0 | 1.4 |
| Asthma | 1986 | 1.0 | 1.4 | 61 | 1.0 | 1.2 |
| Iron deficiency anaemia | 477 | 1.0 | 1.9 | 12 | 0.0 | 0.8 |
| Hypertension | 238 | 1.0 | 1.4 | 5 | 0.0 | 0.4 |
| Other Vaccine Preventable | 57 | 1.0 | 2.2 | <5 | | |
| Pelvic Inflammatory Disease | 126 | 2.0 | 3.1 | <5 | | |
| Nutritional deficiencies | <5 | | | <5 | | |

* rounded to the nearest 5 to avoid disclosure on the number of Nutritional Deficiencies spells.

Table A4. Adult ACSC mortality rates per 1,000 person-years, by ID status and 'main-cause of death' status, 2011-19 (sorted by decreasing main-cause SMR in ID group)

| ACSC | Main-cause | | | | | | All-cause | | | | | |
|---|---|---|---|---|---|---|---|---|---|---|---|---|
| | Crude mortality rate | | | | ID mortality rate age-sex standardized to 'No ID' population | | Crude mortality rate | | | | ID mortality rate age-sex standardized to 'No ID' population | |
| | No ID | | ID | | | | No ID | | ID | | | |
| | CMR | CMR 95% CI | CMR | CMR 95% CI | SMR | SMR 95% CI | CMR | CMR 95% CI | CMR | CMR 95% CI | SMR | SMR 95% CI |
| Any ACSC | 1.03 | (1.00, 1.06) | 1.67 | (1.47, 1.90) | 2.62 | (2.36, 2.91) | 3.90 | (3.84, 3.95) | 5.66 | (5.27, 6.08) | 8.79 | (8.30, 9.30) |
| COPD | 0.80 | (0.78, 0.83) | 0.57 | (0.46, 0.71) | 1.06 | (0.90, 1.25) | 1.73 | (1.70, 1.77) | 1.16 | (1.00, 1.36) | 2.12 | (1.88, 2.38) |
| Convulsions and Epilepsy | 0.02 | (0.02, 0.02) | 0.69 | (0.56, 0.85) | 0.79 | (0.65, 0.95) | 0.10 | (0.09, 0.11) | 2.36 | (2.11, 2.63) | 2.76 | (2.49, 3.06) |
| Asthma | 0.03 | (0.02, 0.03) | 0.10u | (0.06, 0.17) | 0.17u | (0.12, 0.26) | 0.09 | (0.08, 0.10) | 0.23 | (0.16, 0.33) | 0.39 | (0.30, 0.52) |
| Hypertension | 0.03 | (0.03, 0.04) | 0.06u | (0.03, 0.12) | 0.14u | (0.09, 0.22) | 1.15 | (1.12, 1.18) | 0.76 | (0.63, 0.93) | 1.56 | (1.36, 1.78) |
| Congestive Heart Failure | 0.08 | (0.07, 0.09) | 0.07u | (0.04, 0.14) | 0.13u | (0.09, 0.21) | 1.15 | (1.12, 1.18) | 1.25 | (1.08, 1.46) | 2.18 | (1.94, 2.44) |
| Influenza and Pneumonia | 0.03 | (0.02, 0.03) | 0.07u | (0.04, 0.14) | 0.12u | (0.07, 0.20) | 0.04 | (0.03, 0.04) | 0.08u | (0.05, 0.15) | 0.13u | (0.08, 0.20) |
| Cellulitis | 0.02 | (0.02, 0.03) | 0.04u | (0.02, 0.09) | 0.08u | (0.04, 0.15) | 0.05 | (0.04, 0.05) | 0.09u | (0.05, 0.16) | 0.16u | (0.10, 0.24) |
| Pyelonephritis | 0.01 | (0.00, 0.01) | 0.04u | (0.02, 0.09) | 0.06u | (0.03, 0.13) | 0.01 | (0.01, 0.01) | 0.08u | (0.05, 0.15) | 0.12u | (0.07, 0.20) |
| Dehydration and Gastroenteritis | 0.01 | (0.00, 0.01) | | | | | 0.03 | (0.03, 0.04) | 0.12u | (0.07, 0.19) | 0.27u | (0.20, 0.38) |
| Angina | 0.00u | (0.00, 0.00) | | | | | 0.03 | (0.02, 0.03) | | | | |
| Gangrene | | | | | | | 0.02 | (0.01, 0.02) | 0.04u | (0.02, 0.09) | 0.06u | (0.03, 0.13) |
| Pelvic Inflammatory Disease | | | | | | | 0.00u | (0.00, 0.00) | | | | |
| Nutritional Deficiencies | | | | | | | 0.00u | (0.00, 0.00) | | | | |
| Ear, nose and throat infections | | | | | | | 0.00u | (0.00, 0.00) | | | | |
| Dental conditions | | | | | | | 0.00u | (0.00, 0.00) | | | | |
| Iron deficiency anaemia | | | | | | | | | | | | |
| Other Vaccine Preventable | | | | | | | | | | | | |
| Perforated/bleeding ulcer | | | | | | | | | | | | |
| Diabetes Complications | | | | | | | | | | | | |

Note: letter ‘u’ indicates that the figure may not be reliable due to the small number of cases (between 5-20).
