## Supplementary File 1 for "Hospitalisations and deaths due to Ambulatory Care Sensitive Conditions (ACSC) among adults with and without Intellectual Disabilities in Scotland"

### Appendix A

#### Data Sources

- All hospital data are sourced from HES Admitted Patient Care (APC) Commissioning Data Set. It uses published data from April 2013 onwards.
- Organisation Reference Data is sourced from Organisation Data Service (ODS) section in NHS Digital website.

#### ACSC episodes

Finished Consultant Episodes with Admission Method ‘Emergency’ (EPISTAT = 3 and ADMIMETH in '21', '22', '23', '24', '28','2A', '2B', '2C', '2D') where any of the following ICD10 codes are present:

| ACSC name | ICD10 code | Description | Comments |
|---|---|---|---|
| Influenza and pneumonia | J10 | Influenza due to identified influenza virus | In any diagnosis field; exclude people under 3 months or with any secondary diagnosis of D57 |
| | J11 | Influenza, virus not identified | |
| | J13 | Pneumonia due to Streptococcus pneumoniae | |
| | J14 | Pneumonia due to Haemophilus influenzae | |
| | J153 | Pneumonia due to streptococcus, group B | |
| | J154 | Pneumonia due to other streptococci | |
| | J157 | Pneumonia due to Mycoplasma pneumoniae | |
| | J159 | Bacterial pneumonia, unspecified | |
| | J168 | Pneumonia due to other specified infectious organisms | |
| | J181 | Lobar pneumonia, unspecified | |
| | J188 | Other pneumonia, organism unspecified | |
| Other vaccine preventable | A35 | Other tetanus | In any diagnosis field |
| | A36 | Diphtheria | |
| | A37 | Whooping cough | |
| | A80 | Acute poliomyelitis | |
| | B05 | Measles | |
| | B06 | Rubella [German measles] | |
| | B161 | Acute hep B with delta-agent (coinfectn) without hep coma | |
| | B169 | Acute hep B without delta-agent and without hepat coma | |
| | B180 | Chronic viral hepatitis B with delta-agent | |
| | B181 | Chronic viral hepatitis B without delta-agent | |
| | B26 | Mumps | |
| | G000 | Haemophilus meningitis | |
| | M014 | Rubella arthritis | |
| Asthma | J45 | Asthma | Principal diagnosis only |
| | J46 | Status asthmaticus | |

| | | | |
|---|---|---|---|
| Congestive heart failure | I110<br>I50<br>J81 | Hypertensive heart disease with (congestive) heart failure<br>Heart failure<br>Pulmonary oedema | Principal diagnosis only; Exclude main operative procedures with OPCS4 codes of K0,K1,K2,K3,K4,K50,K52,K55,K56,K57, K60,K61,K66,K67,K68,K69,K71 |
| Diabetes complications | E100-E108<br>E110-E118<br>E120-E128<br>E130-E138<br>E140-E148 | Insulin-dependent diabetes mellitus<br>Non-insulin-dependent diabetes mellitus<br>Malnutrition-related diabetes mellitus<br>Other specified diabetes mellitus<br>Unspecified diabetes mellitus | In any diagnosis field |
| Chronic obstructive pulmonary disease | J20<br>J41<br>J42<br>J43<br>J44<br>J47 | Acute bronchitis<br>Simple and mucopurulent chronic bronchitis<br>Unspecified chronic bronchitis<br>Emphysema<br>Other chronic obstructive pulmonary disease<br>Bronchiectasis | Principal diagnosis only; ICD-10: J20 only if there is a secondary diagnosis of J41, J42, J43, J44, J47 |
| Angina | I20<br>I240<br>I248<br>I249 | Angina pectoris<br>Coronary thrombosis not resulting in myocardial infarction<br>Other forms of acute ischaemic heart disease<br>Acute ischaemic heart disease, unspecified | Principal diagnosis only; Exclude cases with main operative procedure OPCS4 codes of A, B,C, D, E, F, G, H, I, J, K, L, M, N, O, P, Q, R, S, T, V, W, X0, X1, X2, X4, X5 |
| Iron deficiency anaemia | D501<br>D508<br>D509 | Sideropenic dysphagia<br>Other iron deficiency anaemias<br>Iron deficiency anaemia, unspecified | Principal diagnosis only |
| Hypertension | I10<br>I119 | Essential (primary) hypertension<br>Hypertensive heart disease without (congestive) heart failure | Principal diagnosis only; Exclude cases with main operative procedure OPCS4 code of K0, K1, K2, K3, K4, K50, K52, K55, K56, K57, K60, K61, K66, K67, K68, K69, K71 |
| Nutritional deficiencies | E40<br>E41<br>E42<br>E43<br>E550<br>E643 | Kwashiorkor<br>Nutritional marasmus<br>Marasmic kwashiorkor<br>Unspecified severe protein-energy malnutrition<br>Rickets, active<br>Sequelae of rickets | Principal diagnosis only |
| Dehydration and gastroenteritis | E86<br>K522<br>K528<br>K529 | Volume depletion<br>Allergic and dietetic gastroenteritis and colitis<br>Other specified noninfective gastroenteritis and colitis<br>Noninfective gastroenteritis and colitis, unspecified | Principal diagnosis only |
| Pyelonephritis | N10<br>N11<br>N12<br>N136 | Acute tubulo-interstitial nephritis<br>Chronic tubulo-interstitial nephritis<br>Tubulo-interstitial nephritis not spec as acute or chronic<br>Pyonephrosis | Principal diagnosis only |
| Perforated/bleeding ulcer | K250-K252,<br>K254-K256<br>K260-K262<br>K264-K266 | Gastric ulcer<br><br>Duodenal ulcer<br> | Principal diagnosis only |

| | | | |
|---|---|---|---|
| | K270-K272 | Peptic ulcer, site unspecified | |
| | K274-K276 | | |
| | K280-K282 | Gastrojejunal ulcer | |
| | K284-K286 | | |
| Cellulitis | L03 | Cellulitis | Principal diagnosis only. Exclude cases with main operative procedure OPCS codes of A, B, C, D, E, F, G, H, I, J, K, L, M, N, O, P, Q, R, S1, S2, S3, S41, S42, S43, S44, S45, S48, S49, T, V, W, X0, X1, X2, X4, X5. Exclude cases with operative procedure OPCS4 code S47 if there are any other operative procedure codes quoted. |
| | L04 | Acute lymphadenitis | |
| | L080 | Pyoderma | |
| | L088 | Other spec local infections of skin and subcutaneous tissue | |
| | L089 | Local infection of skin and subcutaneous tissue, unspecified | |
| | L88 | Pyoderma gangrenosum | |
| | L980 | Pyogenic granuloma | |
| Pelvic inflammatory disease | N70 | Salpingitis and oophoritis | Principal diagnosis only |
| | N73 | Other female pelvic inflammatory diseases | |
| | N74 | Female pelvic inflammatory disorders in diseases EC | |
| Ear, nose and throat infections | H66 | Suppurative and unspecified otitis media | Principal diagnosis only |
| | H67 | Otitis media in diseases classified elsewhere | |
| | J02 | Acute pharyngitis | |
| | J03 | Acute tonsillitis | |
| | J06 | Acute upper respiratory infections multiple and unsp sites | |
| | J312 | Chronic pharyngitis | |
| Dental conditions | A690 | Necrotizing ulcerative stomatitis | Principal diagnosis only |
| | K02 | Dental caries | |
| | K03 | Other diseases of hard tissues of teeth | |
| | K04 | Diseases of pulp and periapical tissues | |
| | K05 | Gingivitis and periodontal diseases | |
| | K06 | Other disorders of gingiva and edentulous alveolar ridge | |
| | K08 | Other disorders of teeth and supporting structures | |
| | K098 | Other cysts of oral region, not elsewhere classified | |
| | K099 | Cyst of oral region, unspecified | |
| | K12 | Stomatitis and related lesions | |
| | K13 | Other diseases of lip and oral mucosa | |
| Convulsions and epilepsy | G40 | Epilepsy | Principal diagnosis only |
| | G41 | Status epilepticus | |
| | R56 | Convulsions, not elsewhere classified | |
| | O15 | Eclampsia | |
| Gangrene | R02 | Gangrene, not elsewhere classified | In any diagnosis field |

#### Classifications

There are slight differences in the way the ACSC are classified. We use the following which is based on a report published by The Health Foundation and Nuffield Trust[1].

| Category | Condition |
|---|---|
| Acute | Cellulitis |
| | Dehydration and gastroenteritis |
| | Dental conditions |
| | Ear, nose and throat infections |
| | Gangrene |
| | Nutritional deficiencies |
| | Pelvic inflammatory disease |
| | Perforated/bleeding ulcer |
| | Pyelonephritis |
| Chronic | Angina |
| | Asthma |
| | Chronic obstructive pulmonary disease |
| | Congestive heart failure |
| | Convulsions and epilepsy |
| | Diabetes complications |
| | Hypertension |
| | Iron deficiency anaemia |
| Vaccine-preventable | Influenza and pneumonia |
| | Other vaccine preventable |

#### Spells

The unit of analysis for ACSC in this report is a continuous inpatient (CIP) spell. A CIP spell is a continuous period of care in hospital, regardless of any transfers which may take place; it may include more than one episode or provider. An episode is a continuous period of admitted patient care under one consultant within one healthcare provider. The length of stay (LoS) is the duration of a spell, i.e. days between admission and discharge. ACSC spells are those CIP spells where the first episode is an ACSC episode.

The method of discharge (DISMETH) is 1, 2 or 3 (Discharged on clinical advice, Self-discharged, or discharged by a mental health review tribunal, i.e. not died, a baby or still in hospital).

For the first episode of the CIP spell (EPIORDER=1):

- The patient classification is an ordinary admission only (CLASSPAT=1).
- The primary diagnosis doesn't relate to Obstetrics (DIAG_01 doesn't begin with 'O')
- It is a General episode (EPITYPE = 1)

[1] Ian Blunt. (2013). Focus on preventable admissions, Tends in emergency admissions for ambulatory care sensitive conditions, 2001 to 2013. *Quality Watch.* Retrieved from https://www.health.org.uk/publications/qualitywatch-focus-on-preventable-admissions on March 2019.

- The age at start of episode is between 0 and 120 (STARTAGE)
- Patient date of birth not 01/01/1900 or 01/01/1901 representing unknown (MYDOB)
- The sex of patient is male or female (SEX is 1 or 2)
- The Main specialty is not

For the last episode of the CIP spell:

- It is a general episode only (EPITYPE=1)
- The Main specialty is not Obstetrics, Midwifery or General Practice with Maternity Function (MAINSPEF) is not 501, 560 or 610

And CIP spells that belong to patients who have had cancer episodes or chemotherapy for cancer both within the reporting year or anywhere in the 365 days prior to admission are not considered ACSC spells (DIAG_01 – DIAG_20 not C00-C97, D37-D48 or Z51.1.

One ACSC condition is assigned to each spell. Conditions numbered 1 to15 in the list below use primary diagnosis at time of admission, so these conditions were assigned to the ACSC spells first. The last four conditions in the list can appear in any diagnosis position in the admission episode and therefore were assigned after the first 15, as only one ACSC condition has been assigned to a spell.

1. Asthma
2. Congestive heart failure
3. Chronic obstructive pulmonary disease
4. Angina
5. Iron deficiency anaemia
6. Hypertension
7. Nutritional deficiencies
8. Dehydration and gastroenteritis
9. Pyelonephritis
10. Perforated/bleeding ulcer
11. Cellulitis
12. Pelvic inflammatory disease
13. Ear, nose and throat infections
14. Dental conditions
15. Convulsions and epilepsy
16. *Gangrene*
17. *Diabetes complications*
18. *Influenza and pneumonia*
19. *Other vaccine preventable*

#### Readmissions

For purposes of this report, if a patient is admitted to hospital within 29 days from the previous discharge, this is considered a readmission. The previous admission must be an ACSC spell for the subsequent admission to be considered as an ACSC readmission. The number of days between discharge date (DISDATE) of the ACSC spell and the admission date (ADMIDATE) of the subsequent admission, irrespective of whether it is an ACSC spell or not, must be between 0 and 29 days for it to be considered a readmission.

### Ethnicity

The ethnicity of the patient, as specified by the patient is recorded using 18 possible values as defined in the 2001 census. In this report they have been grouped as shown below.

| Ethnicity | Ethnicity group |
|---|---|
| A = British (White) | White |
| B = Irish (White) | White |
| C = Any other White background | White |
| D = White and Black Caribbean (Mixed) | Mixed |
| E = White and Black African (Mixed) | Mixed |
| F = White and Asian (Mixed) | Mixed |
| G = Any other Mixed background | Mixed |
| H = Indian (Asian or Asian British) | Asian or Asian British |
| J = Pakistani (Asian or Asian British) | Asian or Asian British |
| K = Bangladeshi (Asian or Asian British) | Asian or Asian British |
| L = Any other Asian background | Asian or Asian British |
| M = Caribbean (Black or Black British) | Black or Black British |
| N = African (Black or Black British) | Black or Black British |
| P = Any other Black background | Black or Black British |
| R = Chinese (other ethnic group) | Other |
| S = Any other ethnic group | Other |
| Z = Not stated | Not stated |
| 99 = Not known (2013 onwards) | Not known |

### Organisation reference data

The Clinical Commissioning Group (CCG) used in this analysis is derived in HES. It contains the code for CCG of Responsibility. In HES, this is derived from postcode of patient’s GP practice, but if that data is not available it uses postcode of patient’s residence and if that data is not available it uses postcode of the provider to lookup the CCG code from the Postcode Directory which is available in ODS section in the NHS Digital website.

The Sustainability and Transformation Partnership (STP) codes are derived using reference data available in ODS section in NHS Digital website from CCG of responsibility present in HES.

### Suppression

Data suppression is based on HES Disclosure Control Methodology for Hospital Episode Statistics for the Admitted Patient Care (APC) data set. For the counts of spells: Zeroes remain unchanged, counts between 1 and 7 (inclusive) are shown as 5 and all other counts are rounded to the nearest 5.
